# SARS-CoV-2 transmission in K-12 schools in the Vancouver Coastal Health Region: a descriptive epidemiologic study

**DOI:** 10.1101/2021.05.15.21257271

**Authors:** Diana Bark, Nalin Dhillon, Martin St-Jean, Brooke Kinniburgh, Geoff McKee, Alexandra Choi

## Abstract

**Background:** There is an urgent need to assess the role of schools in the spread of SARS-CoV-2 in Canada to inform public health measures. We describe the epidemiology of SARS-CoV-2 infection in students and staff in the Vancouver Coastal Health (VCH) region in the first three months of the 2020/2021 academic year, and examine the extent of transmission in schools.

**Methods:** This descriptive epidemiologic study using contact tracing data included all SARS-CoV-2 cases reported to VCH between September 10 and December 18, 2020 who worked in or attended K-12 schools in-person. Case and cluster characteristics were described.

**Results:** There were 699 school staff and student cases during the study period, for an incidence of 55 cases per 10,000 population, compared to 73 per 10,000 population in all VCH residents. Among VCH resident staff and student cases, 53% were linked to a household case/cluster, <1.5% were hospitalized and there were no deaths. Out of 699 cases present at school, 26 clusters with school-based transmission resulted in 55 secondary cases. Staff members accounted for 54% of index cases (14/26) while comprising 14% of the school population. Among clusters, 88% had fewer than 4 secondary cases.

**Interpretation:** COVID-19 incidence in the school population was lower than that of the general population. There were no deaths and severe disease was rare. School-based transmissions of SARS-CoV-2 were uncommon and clusters were small. Our results support the growing body of evidence that schools do not play a major role in the spread of SARS-CoV-2.

## Introduction

Early in the pandemic, schools closed across Canada to help reduce the circulation of SARS-CoV-2 and decrease risks associated with infection among staff and students(1). Evidence has since suggested that school closures and decreased in-person learning have had negative impacts on the mental health, wellbeing, educational attainment, social connection, healthcare access, and health behaviors of students, families, and staff(1-12). Combined with high socioeconomic tolls(13-15), these unintended consequences have compelled consideration of the efficacy of school closures in limiting morbidity and mortality associated with the COVID-19 pandemic.

Evidence now indicates that age is a risk factor for severe disease and children typically experience a mild course of illness(16-22). There is also a growing body of literature suggesting children’s in-person school attendance may not be a significant driver of transmission; contact-tracing and test-based studies from other jurisdictions have identified little or no transmission in the school setting(23-34). Internationally, studies illustrate that school re-opening has not impacted numbers of paediatric cases(23,35,36) and that the risk of SARS-CoV-2 infection has not increased for educational staff working with children(30,36,37). There is limited peer-reviewed data in Canada regarding risk of SARS-COV-2 transmission in schools, although Ottawa Public Health has reported low rates of school-based transmission(38). Given that transmission risk is influenced by community-level epidemiology, public health measures, health system infrastructure, and other jurisdictional factors, this study contributes needed evidence within the Canadian context. We aimed to describe cases who attended kindergarten to grade 12 (K-12) schools in-person, assess the extent of SARS-CoV-2 transmission in schools, and characterize clusters where school-based transmission was observed.

## Methods

### Setting

During the study period, while mass gatherings remained prohibited, schools and businesses were open with safety plans in place, and masks were encouraged but not required. Travel and gatherings in private residences were restricted gradually starting in November 2020. There were no stay-at-home requirements(39). Symptomatic testing was widely available and contact tracing was conducted for all positive cases(40).

Vancouver Coastal Health (VCH) serves over 1.1 million people(41) across urban, rural, and remote geographies. During the study period, 17,742 staff and 105,905 students worked in or attended school in-person at a total of 358 schools (104 independent and 254 public) in the VCH region. Schools in BC reopened following summer holidays on September 10, 2020 and closed for winter break after December 18, 2020. Prior to reopening, schools implemented COVID-19 safety plans developed with support from Public Health, which included public health measures (e.g. testing and contact tracing protocols), environmental measures (e.g. enhanced cleaning and disinfection), administrative measures (e.g. scheduling and work practices, health and wellness policies, and cohorting), personal measures (e.g. staying home when sick, physical distancing, hand hygiene and respiratory etiquette), and personal protective equipment. Non-medical mask use was supported, but not required(42).

### Public Health Investigation and Response

SARS-CoV-2 Nucleic Acid Amplification testing (NAAT) was available for anyone with symptoms, and advised for students or staff with fever or new symptoms persisting >24 hours. Tests were generally processed within 24 hours. Positive tests were automatically reported to VCH’s Office of the Chief Medical Health Officer, which investigated cases within 24 hours and identified contacts(43). Individual risk assessments were performed for all contacts, integrating cases’ symptoms, ages of cases and contacts, nature and duration of contact, setting (e.g. indoor/outdoor), and presence/absence of known SARS-CoV-2 transmission. Factors constituting high-risk exposures included direct contact with infectious body fluids (e.g., via sharing vapes, being coughed or sneezed upon) and prolonged (>15 minutes) contact with a symptomatic case face to face or at close range; thus, close contacts were identified among classmates. Factors constituting low-risk exposures included the absence of interactions or interactions outdoors of limited duration and at a distance (>2m). Individuals with high-risk exposures were advised to self-isolate for 14 days and were monitored by Public Health for emergence of symptoms during their incubation period. Symptomatic individuals were directed to seek testing and few declined. Those with lower risk exposures were advised to self-monitor for symptoms and present for testing accordingly. Asymptomatic individuals were tested at the discretion of the Medical Health Officer. Mass asymptomatic testing was undertaken in outbreak settings where uncontrolled spread was observed; this did not occur in any schools during the study period, and there were no school closures due to SARS-CoV-2 transmission.

### Data sources

We obtained data from VCH’s electronic COVID-19 case and contact management platform, capturing contact tracing interview data including demographics, school name, grade, role (staff/student), attendance dates, comorbidities, symptoms, and epidemiologic linkage to other cases within and outside of school. Hospitalizations and ICU admissions to VCH hospitals were determined using individual data linkages to acute care data. Provincial Health Services Authority provided hospitalization data for BC Children’s Hospital. Districts confirmed denominators of staff and students attending in-person during the study period. Duplicate case entries were discarded based on unique identifiers. Cases with missing or incorrect locations were reclassified based on city of residence or postal code.

### Study design

We extracted all lab-confirmed and probable cases aged 5 and older at time of case report between September 10 and December 18, 2020 who reported working in or attending a K-12 school within VCH during their incubation or infectious periods. We excluded cases who exclusively worked in or attended a daycare or post-secondary institution, attended school online, or were homeschooled. School staff and student cases were categorized as either school aged (aged 5-17) or aged 18+. We described case characteristics including age, sex, location, linkage to known cases/clusters, comorbidities, symptoms at time of initial interview, status (recovered/removed from isolation or deceased), and hospitalization/ICU admission.

To identify potential school transmission events, we analyzed all clusters where 2 or more cases were reported within a 14-day period in a school. Cases who did not interact with staff or students in the learning setting (e.g., building maintenance staff only present after school hours) were excluded. A chart review of the cluster cases and their epidemiologically linked cases was conducted to verify the likely source of SARS-CoV-2 infection. A case was determined to be a contact’s likely source if they had prolonged contact during the case’s infectious period and the contact’s incubation period. Sources were categorized as household, social, school, or other. Index and secondary cases were determined by symptom onset date. If there was no contact with a positive case, this was recorded. All cases with multiple potential exposures were reviewed by DB and AC. The source of acquisition was attributed to the school if the non-school source had an unclear symptom onset date and no clear source of acquisition themselves. Finally, secondary cases’ close contacts who became lab-confirmed or probable cases were deemed tertiary cases. The primary outcome was the number of school-based transmissions.

All analyses were conducted via statistical computing software R Studio(44). P-values were based on Pearson’s chi-squared and independent t-tests for categorical and continuous variables respectively.

## Results

### Description of cases in schools

In the four weeks prior to school reopening, weekly incidence of SARS-CoV-2 infection ranged 0.8-3.2 per 10,000 population in VCH based on 2019 population estimates(41), compared to 2.4-2.8 per 10,000 population in the four weeks following reopening. Weekly incidence peaked November 15-21 among VCH residents at 9.4 per 10,000 population and among K-12 staff and students at 6.6 per 10,000 population. Between September 10 and December 18, 8,755 cases among VCH residents were reported (73 per 10,000 population), of which 674 (8%) were K-12 staff or students. With an additional 25 staff and student cases who resided outside the VCH region, there were 699 cases among 123,646 staff and students working in or attending school in-person (55 per 10,000 population) in 270 unique VCH region K-12 schools during the same period. Characteristics of the 674 VCH resident staff and student cases are described in Table 1. Notably, adults (18+) more frequently reported comorbidities and were less frequently linked to a confirmed case/cluster compared to children (5-17 years) (p-values <0.01).

**Figures 1A and B.**
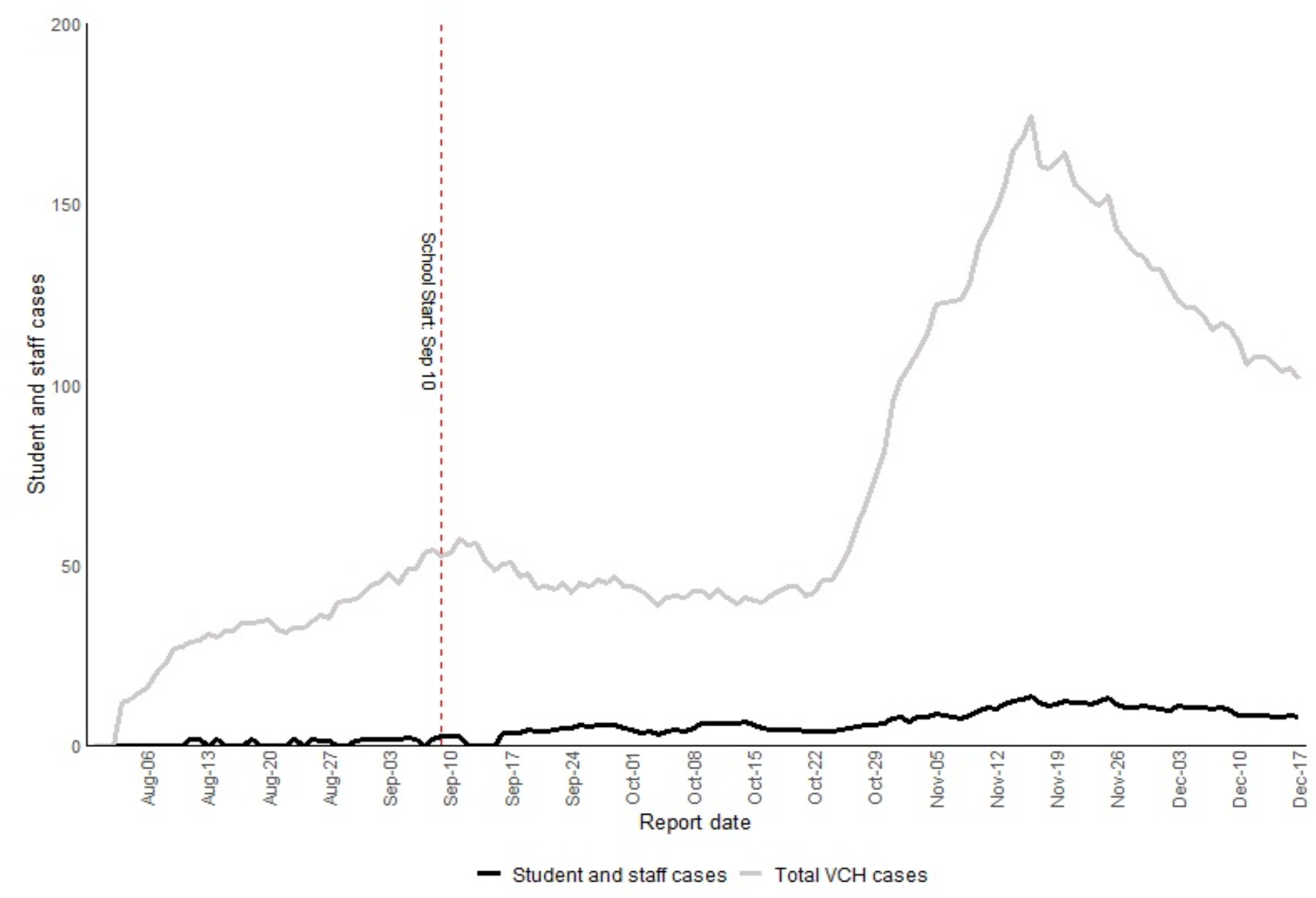
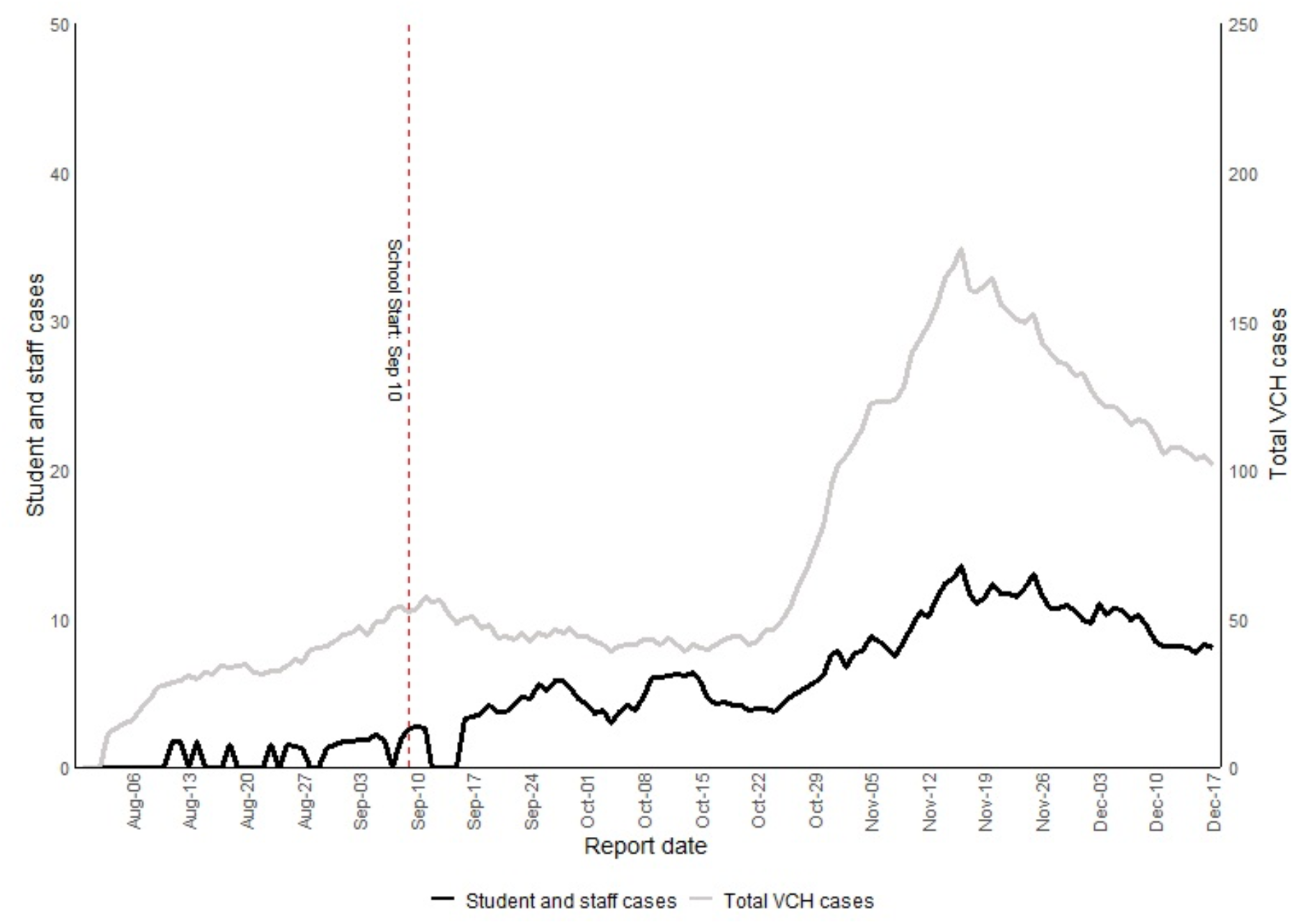
**7-day average of new SARS-CoV-2 cases among VCH residents, reported between August 1 and December 18, 2020. *Please note the different axis used in Figure 1b to compare trends between school and community cases***

**Table 1.**
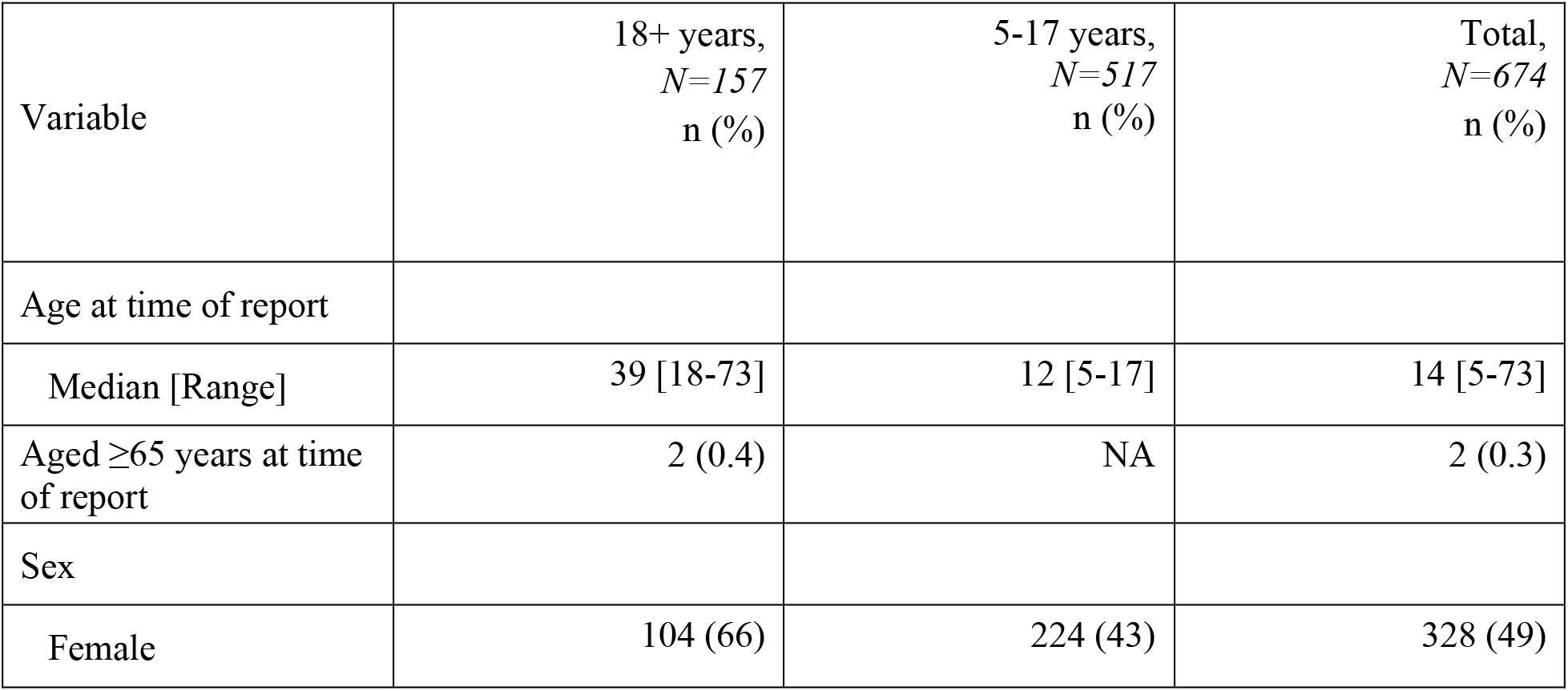
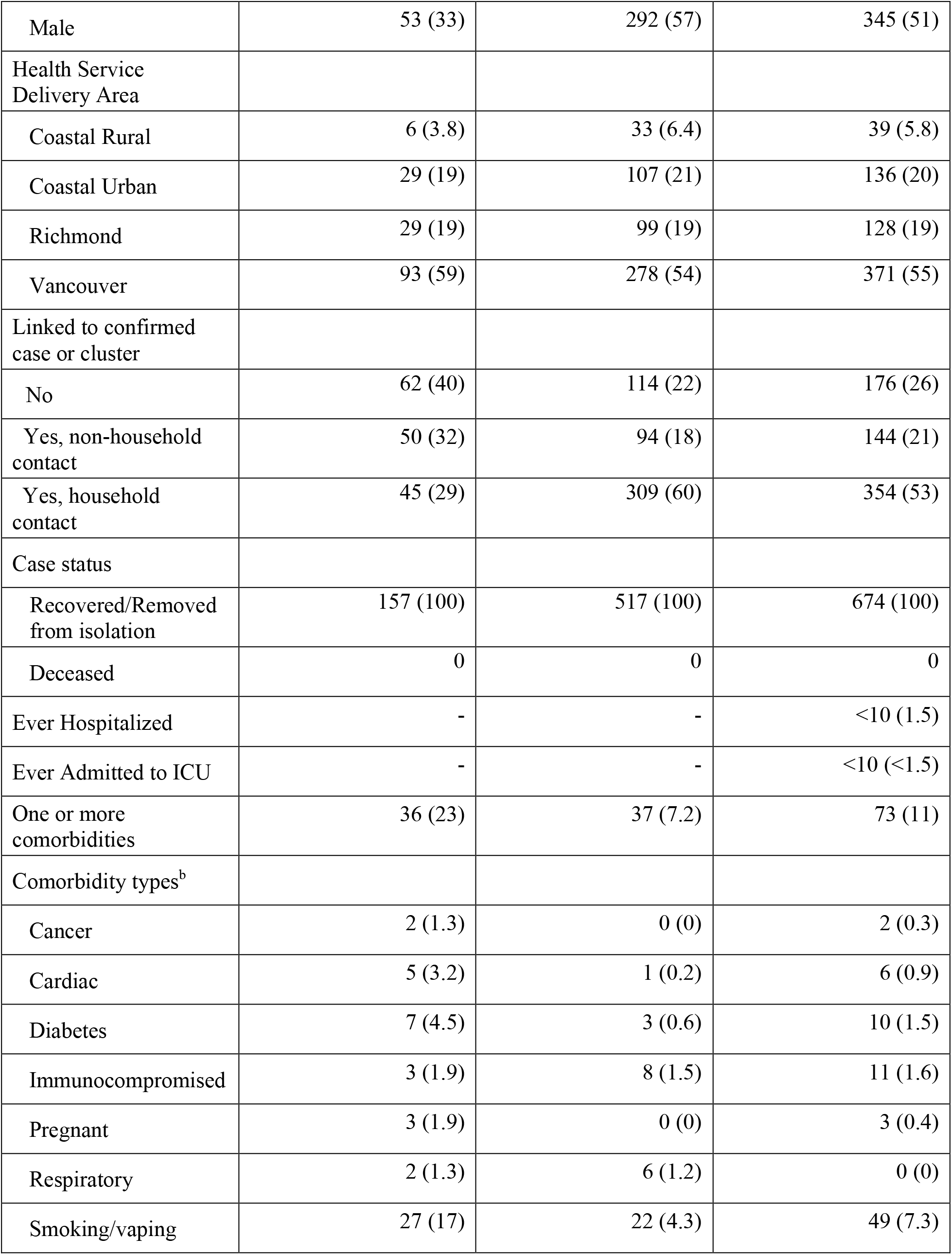
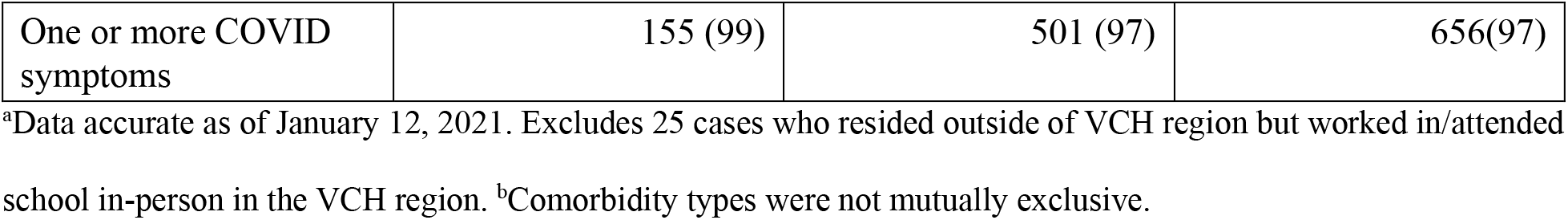
**Characteristics of 674 student and staff SARS-CoV-2 cases residing in the VCH region reported between September 10 and December 18, 2020**^**a**^

| Variable | 18+ years,<br>*N*=157<br>*n* (%) | 5-17 years,<br>*N*=517<br>*n* (%) | Total,<br>*N*=674<br>*n* (%) |
| --- | --- | --- | --- |
| Age at time of report |  |  |  |
| Median [Range] | 39 [18-73] | 12 [5-17] | 14 [5-73] |
| Aged ≥65 years at time of report | 2 (0.4) | NA | 2 (0.3) |
| Sex |  |  |  |
| Female | 104 (66) | 224 (43) | 328 (49) |
| Male | 53 (33) | 292 (57) | 345 (51) |
| Health Service Delivery Area |  |  |  |
| Coastal Rural | 6 (3.8) | 33 (6.4) | 39 (5.8) |
| Coastal Urban | 29 (19) | 107 (21) | 136 (20) |
| Richmond | 29 (19) | 99 (19) | 128 (19) |
| Vancouver | 93 (59) | 278 (54) | 371 (55) |
| Linked to confirmed case or cluster |  |  |  |
| No | 62 (40) | 114 (22) | 176 (26) |
| Yes, non-household contact | 50 (32) | 94 (18) | 144 (21) |
| Yes, household contact | 45 (29) | 309 (60) | 354 (53) |
| Case status |  |  |  |
| Recovered/Removed from isolation | 157 (100) | 517 (100) | 674 (100) |
| Deceased | 0 | 0 | 0 |
| Ever Hospitalized | - | - | <10 (1.5) |
| Ever Admitted to ICU | - | - | <10 (<1.5) |
| One or more comorbidities | 36 (23) | 37 (7.2) | 73 (11) |
| Comorbidity types<sup>b</sup> |  |  |  |
| Cancer | 2 (1.3) | 0 (0) | 2 (0.3) |
| Cardiac | 5 (3.2) | 1 (0.2) | 6 (0.9) |
| Diabetes | 7 (4.5) | 3 (0.6) | 10 (1.5) |
| Immunocompromised | 3 (1.9) | 8 (1.5) | 11 (1.6) |
| Pregnant | 3 (1.9) | 0 (0) | 3 (0.4) |
| Respiratory | 2 (1.3) | 6 (1.2) | 0 (0) |
| Smoking/vaping | 27 (17) | 22 (4.3) | 49 (7.3) |
| One or more COVID symptoms | 155 (99) | 501 (97) | 656(97) |

<sup>a</sup>Data accurate as of January 12, 2021. Excludes 25 cases who resided outside of VCH region but worked in/attended school in-person in the VCH region. <sup>b</sup>Comorbidity types were not mutually exclusive.

### Clusters of SARS-CoV-2 infection in schools

There were 71 school clusters totaling 251 cases during the study period. No school-based transmission was suspected in 45 clusters (63%), with all 142 cases within these clusters likely acquiring SARS-CoV-2 outside of school. Within the 26 clusters containing at least one school-acquired case, a further 28 cases likely acquired infection outside of school. Of the 170 cases (19 staff and 151 students) temporally clustered but without evidence of school-based transmission, 107 (63%) were attributable to household contacts, 17 (10%) to social networks, 4 (2.4%) to sports teams, and 2 (1.2%) to work outside of school. 40 students and staff (24%) had no known contact with another case inside or outside of school.

### Cases with evidence of school-based transmission

Out of 699 cases present at school, transmission stemming from 26 index cases resulted in 55 secondary cases at school. Two hundred and thirty-seven close contacts were identified from these 55 secondary cases, and 10 became tertiary cases. Staff accounted for 54% of index cases (14/26) and 25% of secondary cases (14/55). Staff cases (n=28) included 19 teachers, 6 support workers, and 3 principals/office staff members. Of the 81 index and secondary cases, 47 (58%) worked in or attended a K-7 school, 25 (31%) a grade 8-12 school, and 9 (11%) a K-12 school.

The characteristics of clusters with school-based transmission are presented in Table 2. Among 26 total clusters, 13 (50%) had only 1 secondary case, 10 (38%) had 2-3 secondary cases, and 3 (12%) had 4 or more secondary cases. All clusters with 4 or more secondary cases had an index staff case and occurred in grades K-7. The median size of clusters was 3 (range 2-8) when the index case was a staff member compared to 2 (range 2-3) when the index case was a student. There were 8 (31%) clusters involving students only, all among Grades 4-12, and 4 clusters (15%) involving staff members only. Of the 14 clusters including both staff and students, 9 (64%) occurred in grades K-7 and 5 (36%) in grades 8-12.

**Table 2.**
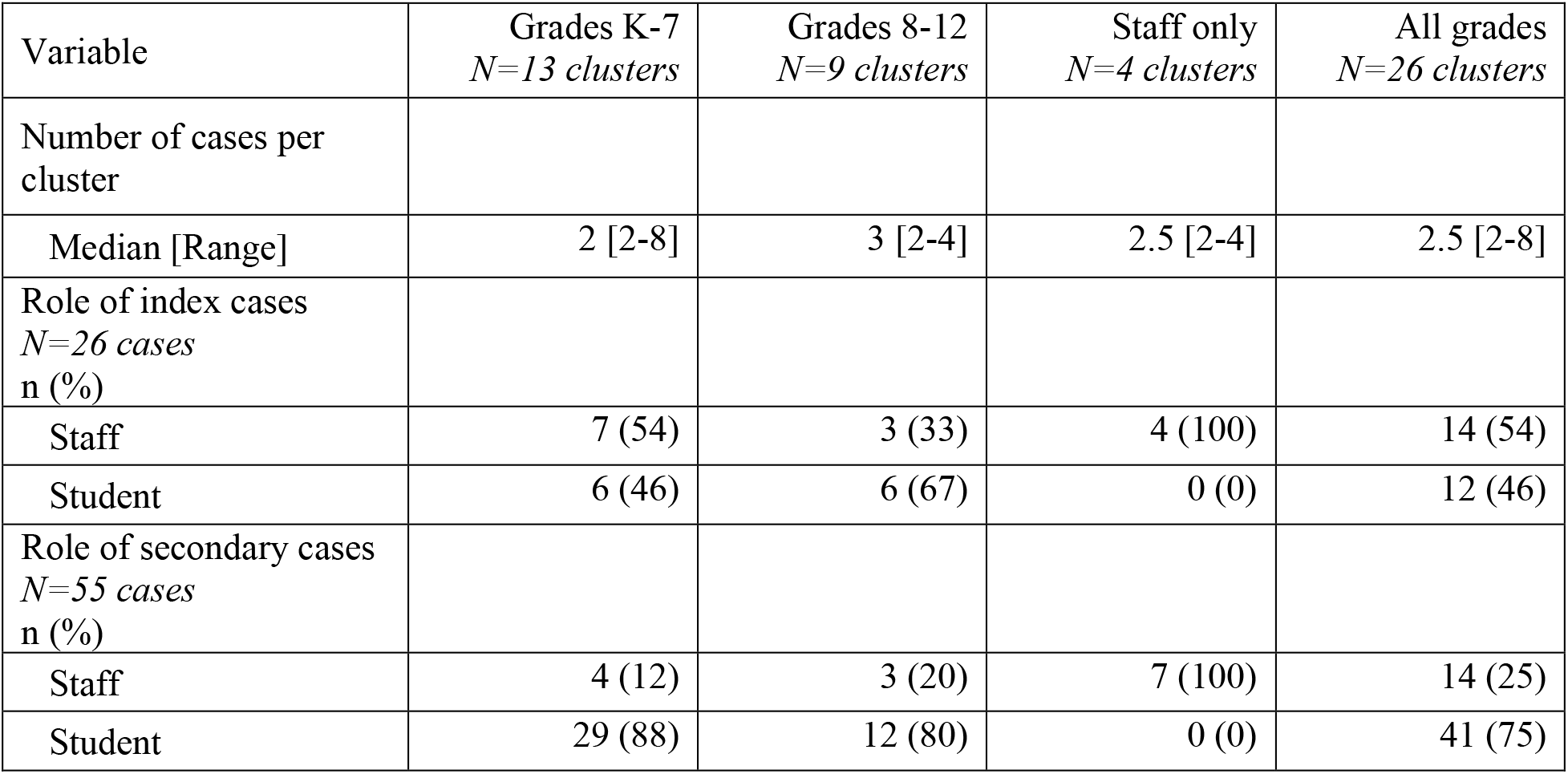
Characteristics of 26 clusters with evidence of school-based transmission.

School-based activities where transmission may have occurred are presented in Table 3. Evidence of transmission was most commonly found in a shared classroom, but no specific class type was more frequently identified. For some cases, there were multiple exposures settings within the school, notably through socializing with a classmate.

**Table 3.** **School settings where transmission may have occurred for 55 school-acquired SARS-CoV-2 cases**^**a**^

| Exposure setting | Grades K-7<br>*N=35 exposure settings* | Grades 8-12<br>*N=17 exposure settings* | Staff only<br>*N=7 exposure settings* | All grades<br>n (%)<br>*N=59 exposure settings* |
| --- | --- | --- | --- | --- |
| Class | 28<br>(26 students,<br>2 staff) | 11<br>(10 students,<br>1 staff) | 2<br>(2 staff) | 41 (70) |
| Office/Meeting/Staff room | 2<br>(2 staff) | 1<br>(1 staff) | 3<br>(3 staff) | 6 (10) |
| Socializing at school | 3<br>(3 students) | 2<br>(2 students) | 0 | 5 (8) |
| Unspecified interaction | 0 | 1<br>(1 student) | 2<br>(2 staff) | 3 (5) |
| Field trip | 0 | 2<br>(1 student,<br>1 staff) | 0 | 2 (3) |
| Cohort<sup>b</sup> | 2<br>(2 students) | 0 | 0 | 2 (3) |

<sup>a</sup>There were multiple potential exposure settings for some cases; an assessment of which within-school exposure setting was most likely responsible for transmission was not included as part of this study.
<sup>b</sup>A cohort is a group of students and staff who remain together throughout a school term, the composition of which is consistent for all activities that occur in schools.

## Interpretation

During the study period 8,755 cases of SARS-CoV-2 infection were reported among VCH residents (73 per 10,000 population) compared to 699 among VCH K-12 student and staff (55 per 10,000 population). While 251 cases were temporally clustered with another case in the same school within a 14-day period, 170 (68%) of these cases had no evidence of school-based transmission. We identified 26 clusters with evidence of school-based transmission resulting in 55 school-acquired cases (0.04% of 123,646 individuals working in or attending school) in 25/358 (7.0%) K-12 schools in the VCH region. Cluster sizes were small, with 50% leading to 1 secondary case and 88% having fewer than 5 cases total; largest clusters were observed in elementary schools. The index case was a staff member in over half of the clusters (54%), although staff only constituted 35% (28/81) of all cases linked to school-based transmission and 14% of the overall school community. A shared classroom accounted for most transmissions; no particular class/activity was noticeably higher risk.

Our findings support the growing body of evidence that schools are likely not a significant driver of SARS-CoV-2 transmission(23,35,36). In concordance with other studies, we found that a household case was the predominant source of infection for school-aged children(30,32), and that school-based acquisitions were uncommon(29,30,32,33). Individuals with COVID-19 seldom infected others at school; out of 699 student and staff cases, we identified school-based transmissions stemming from 26 index cases (3.9%). The ratio of secondary cases to total primary cases was 0.09 in the school setting, compared to 0.27 observed in Luxembourg(30), 0.08 in Germany(32) and 0.02 in South Korea(23). While we did not systematically track the number of school contacts per index case, the secondary attack rate is likely to be very low given the low number of identified secondary cases, in concordance with prior studies across several countries(24,25,28,30). As observed elsewhere, where school-based transmission occurred, the number of secondary cases was limited(23,31,45), although one large high school outbreak was described in Israel(46) in the context of crowding and a concurrent heat wave. Interestingly, we observed a larger number of secondary cases when the index case was an adult, similar to the experience in England(31) and Singapore(26). Finally, our findings reinforced the observation that numerous cases in a school can reflect multiple introductions from the community rather than school-based transmissions(2).

Of note, our study was conducted in the context of limited circulation of SARS-CoV-2 variants of concern, strong collaboration between the public health and education sectors, good school-based infection prevention and control measures, and rapid access to testing and contact tracing. Contact tracing within 24 hours and regular monitoring of cases and contacts may have prevented transmission to tertiary cases. Further studies will be required to evaluate the impact of individual infection prevention and control measures.

## Limitations

Our study was population-based and avoided bias due to cluster selection. However, our analysis was limited by reliance on routinely collected data and by potential recall bias, although case/proxy report was supplemented with information from school administrators as needed. While Medical Health Officers could request asymptomatic testing for the purposes of public health investigation, universal testing was not required. Transmissions to undetected (e.g. asymptomatic and not tested) secondary and tertiary cases were therefore not examined, which could disproportionately affect groups more likely to experience asymptomatic infection. However, studies that employed asymptomatic testing of school contacts reported low levels of transmission(24,25,28). Class sizes were not measured; thus, any association with school-based transmissions was not assessed. The potential for misclassification of the likely source of acquisition for cases with multiple exposures was minimized via independent review of these cases by two members of the research team. This study was limited to descriptive statistics and causal inferences cannot be drawn.

## Conclusion

In conclusion, from September 10 to December 18, 2020 we detected minimal clusters and low rates of secondary transmission at schools. In-person school attendance may not expose students and staff to higher risks than those experienced in the community when infection prevention and control measures are in place, and adequate case and contact management capacity is available. Caution should be taken in generalizing these findings due to differences in underlying epidemiology, public health measures, health system infrastructure, and culture, among other factors that may differ by jurisdictions. However, our findings suggest that schools may be able to safely remain open in settings with moderate community transmission (i.e., weekly incidence of 10-100 cases per 100,000 as observed in this study) with infection prevention and control measures in place and sufficient public health capacity for rapid testing and contact tracing. Acknowledging harms associated with decreased school attendance, policy makers should focus on the implementation of measures to safely operate in-person learning, and consider school closures and widespread quarantine of students and staff with great caution.

## Supporting information

RECORD Checklist

## Data Availability

The study data are not available for use by other researchers, other than that which are presented in the manuscript.

## Acknowledgements

We thank Amina Moustaqim-Barrette MPH (Vancouver Coastal Health) and Moe Zandy MSc (British Columbia Centre for Disease Control) for their contributions to data compilation and visualization.

## Disclaimers

The views expressed in the article are the authors’ own and not an official position of Vancouver Coastal Health or the British Columbia Centre for Disease Control.

## Disclosures

AC is a Medical Health Officer and School Medical Officer for Vancouver Coastal Health. GM is the Medical Director for Population and Public Health at the BC Centre for Disease Control. Neither the study authors nor their institutions received payment or services at any time from any third parties for any aspects of the submitted work.

## Ethical considerations

In consultation with UBC Office of Research Ethics, this study did not require institutional research ethics review according to Article 2.5 of the Tri-Council Policy Statement (TCPS2), as it was completed as part of routine public health surveillance and quality assurance/quality improvement.

## Appendix

The study protocol and BC Centre for Disease Control COVID-19 Public Health Guidance for K-12 School Settings document are available upon request by emailing the corresponding author.

**Table A1.**
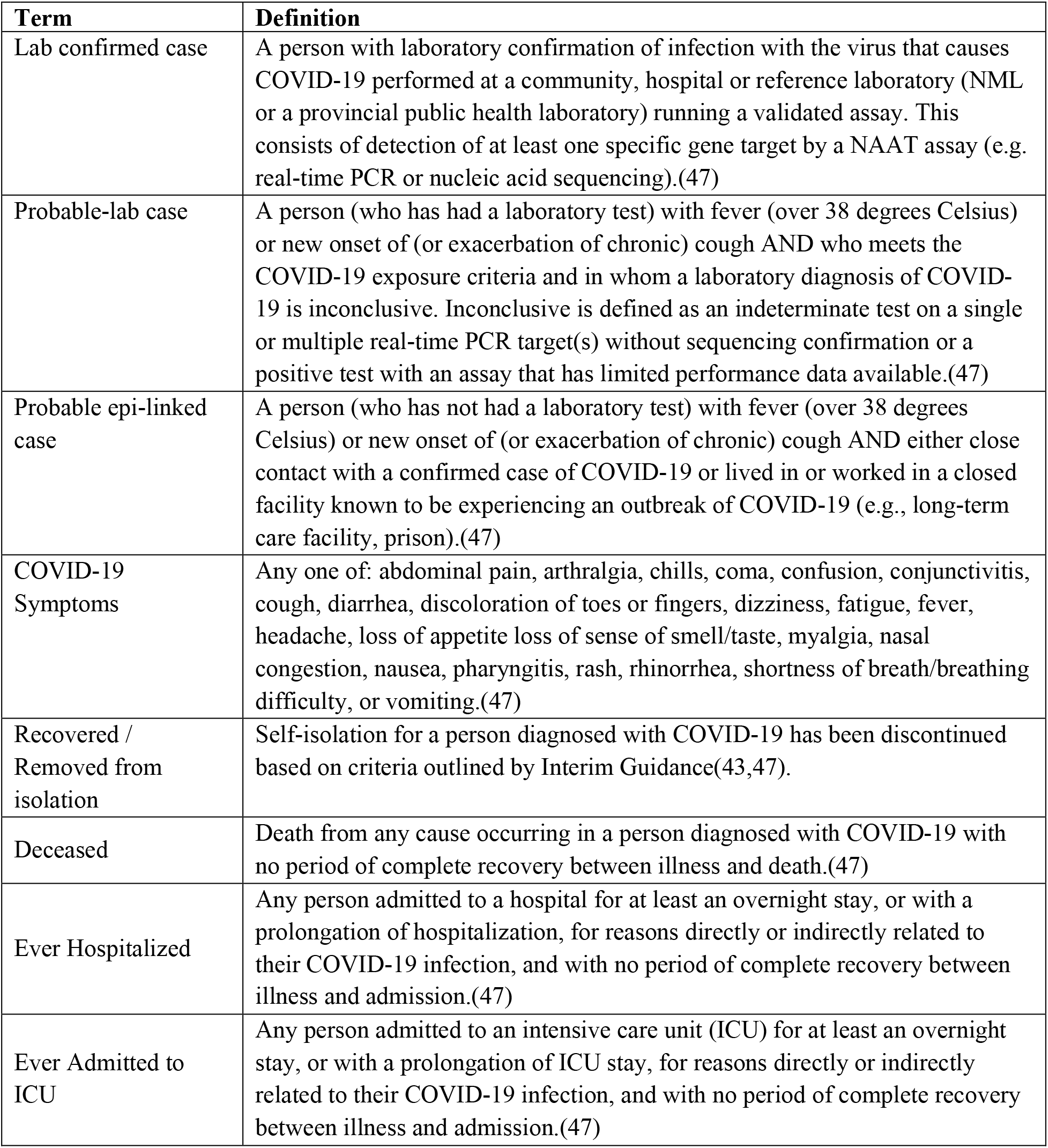
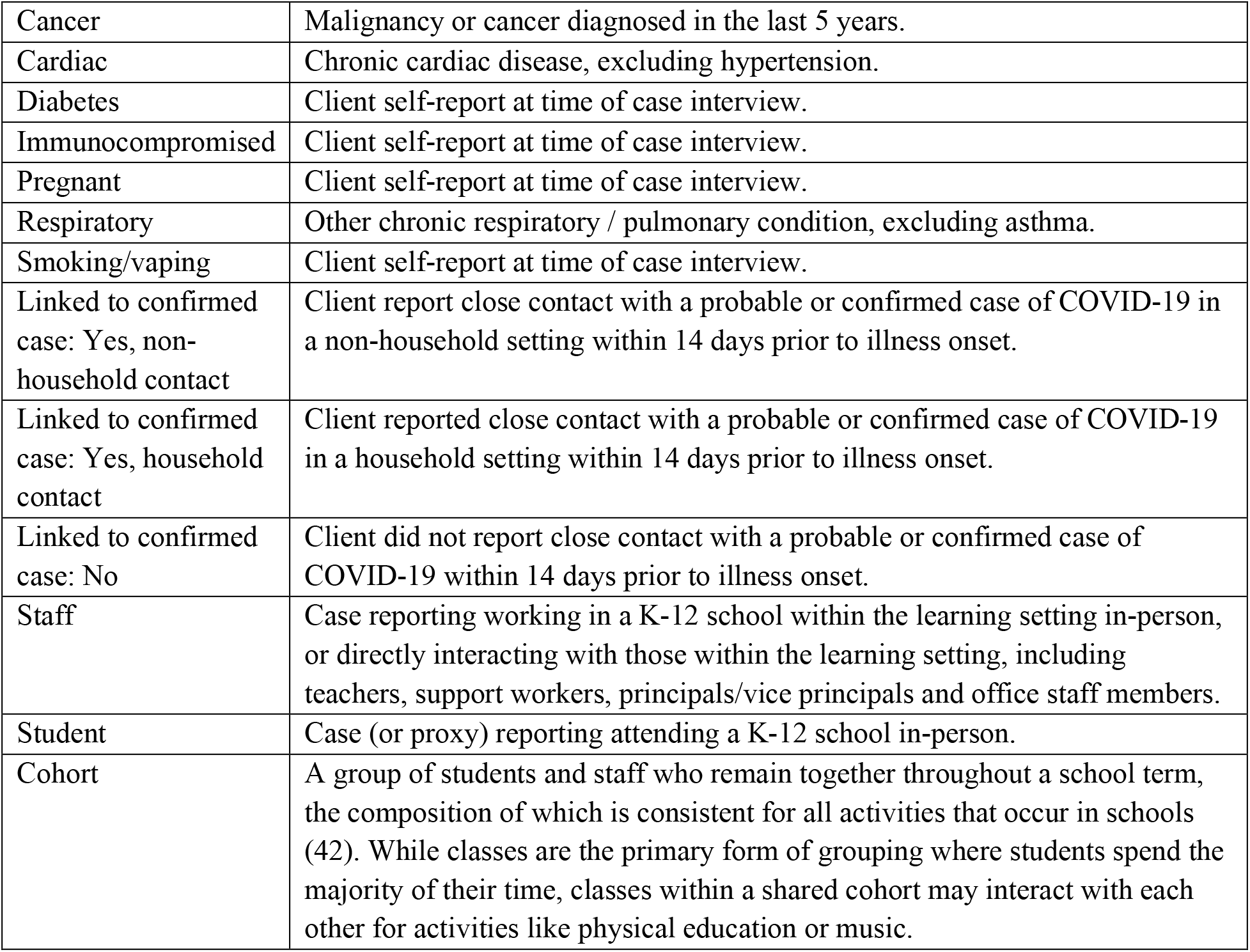
Data dictionary.

| **Term** | **Definition** |
| --- | --- |
| Lab confirmed case | A person with laboratory confirmation of infection with the virus that causes COVID-19 performed at a community, hospital or reference laboratory (NML or a provincial public health laboratory) running a validated assay. This consists of detection of at least one specific gene target by a NAAT assay (e.g. real-time PCR or nucleic acid sequencing).(47) |
| Probable-lab case | A person (who has had a laboratory test) with fever (over 38 degrees Celsius) or new onset of (or exacerbation of chronic) cough AND who meets the COVID-19 exposure criteria and in whom a laboratory diagnosis of COVID-19 is inconclusive. Inconclusive is defined as an indeterminate test on a single or multiple real-time PCR target(s) without sequencing confirmation or a positive test with an assay that has limited performance data available.(47) |
| Probable epi-linked case | A person (who has not had a laboratory test) with fever (over 38 degrees Celsius) or new onset of (or exacerbation of chronic) cough AND either close contact with a confirmed case of COVID-19 or lived in or worked in a closed facility known to be experiencing an outbreak of COVID-19 (e.g., long-term care facility, prison).(47) |
| COVID-19 Symptoms | Any one of: abdominal pain, arthralgia, chills, coma, confusion, conjunctivitis, cough, diarrhea, discoloration of toes or fingers, dizziness, fatigue, fever, headache, loss of appetite loss of sense of smell/taste, myalgia, nasal congestion, nausea, pharyngitis, rash, rhinorrhea, shortness of breath/breathing difficulty, or vomiting.(47) |
| Recovered / Removed from isolation | Self-isolation for a person diagnosed with COVID-19 has been discontinued based on criteria outlined by Interim Guidance(43,47). |
| Deceased | Death from any cause occurring in a person diagnosed with COVID-19 with no period of complete recovery between illness and death.(47) |
| Ever Hospitalized | Any person admitted to a hospital for at least an overnight stay, or with a prolongation of hospitalization, for reasons directly or indirectly related to their COVID-19 infection, and with no period of complete recovery between illness and admission.(47) |
| Ever Admitted to ICU | Any person admitted to an intensive care unit (ICU) for at least an overnight stay, or with a prolongation of ICU stay, for reasons directly or indirectly related to their COVID-19 infection, and with no period of complete recovery between illness and admission.(47) |
| Cancer | Malignancy or cancer diagnosed in the last 5 years. |
| Cardiac | Chronic cardiac disease, excluding hypertension. |
| Diabetes | Client self-report at time of case interview. |
| Immunocompromised | Client self-report at time of case interview. |
| Pregnant | Client self-report at time of case interview. |
| Respiratory | Other chronic respiratory / pulmonary condition, excluding asthma. |
| Smoking/vaping | Client self-report at time of case interview. |
| Linked to confirmed case: Yes, non-household contact | Client report close contact with a probable or confirmed case of COVID-19 in a non-household setting within 14 days prior to illness onset. |
| Linked to confirmed case: Yes, household contact | Client reported close contact with a probable or confirmed case of COVID-19 in a household setting within 14 days prior to illness onset. |
| Linked to confirmed case: No | Client did not report close contact with a probable or confirmed case of COVID-19 within 14 days prior to illness onset. |
| Staff | Case reporting working in a K-12 school within the learning setting in-person, or directly interacting with those within the learning setting, including teachers, support workers, principals/vice principals and office staff members. |
| Student | Case (or proxy) reporting attending a K-12 school in-person. |
| Cohort | A group of students and staff who remain together throughout a school term, the composition of which is consistent for all activities that occur in schools (42). While classes are the primary form of grouping where students spend the majority of their time, classes within a shared cohort may interact with each other for activities like physical education or music. |

**Figure A1.**
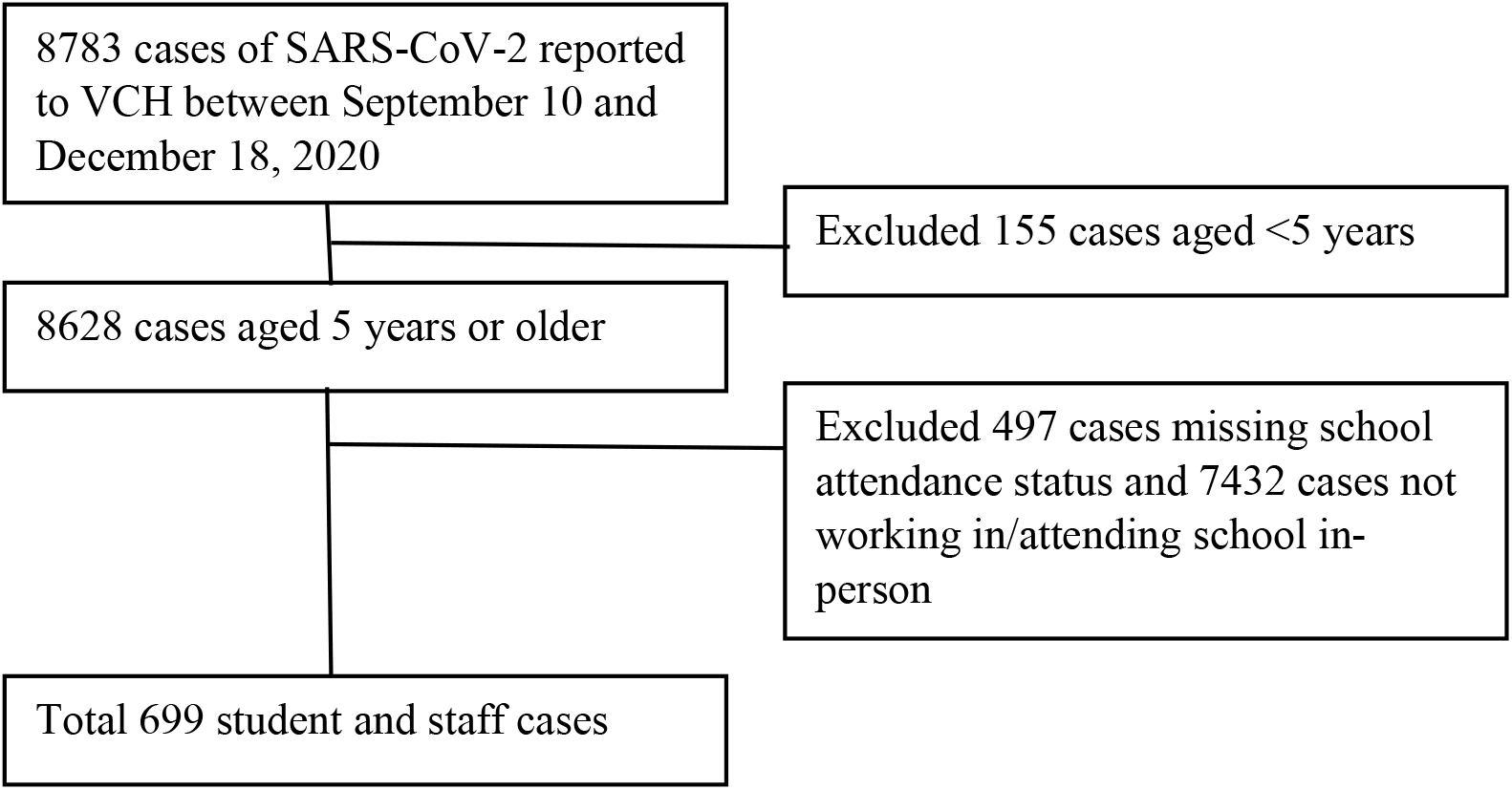
Study population selection flow diagram (including non-VCH resident staff and student cases)

**Figure A2.**
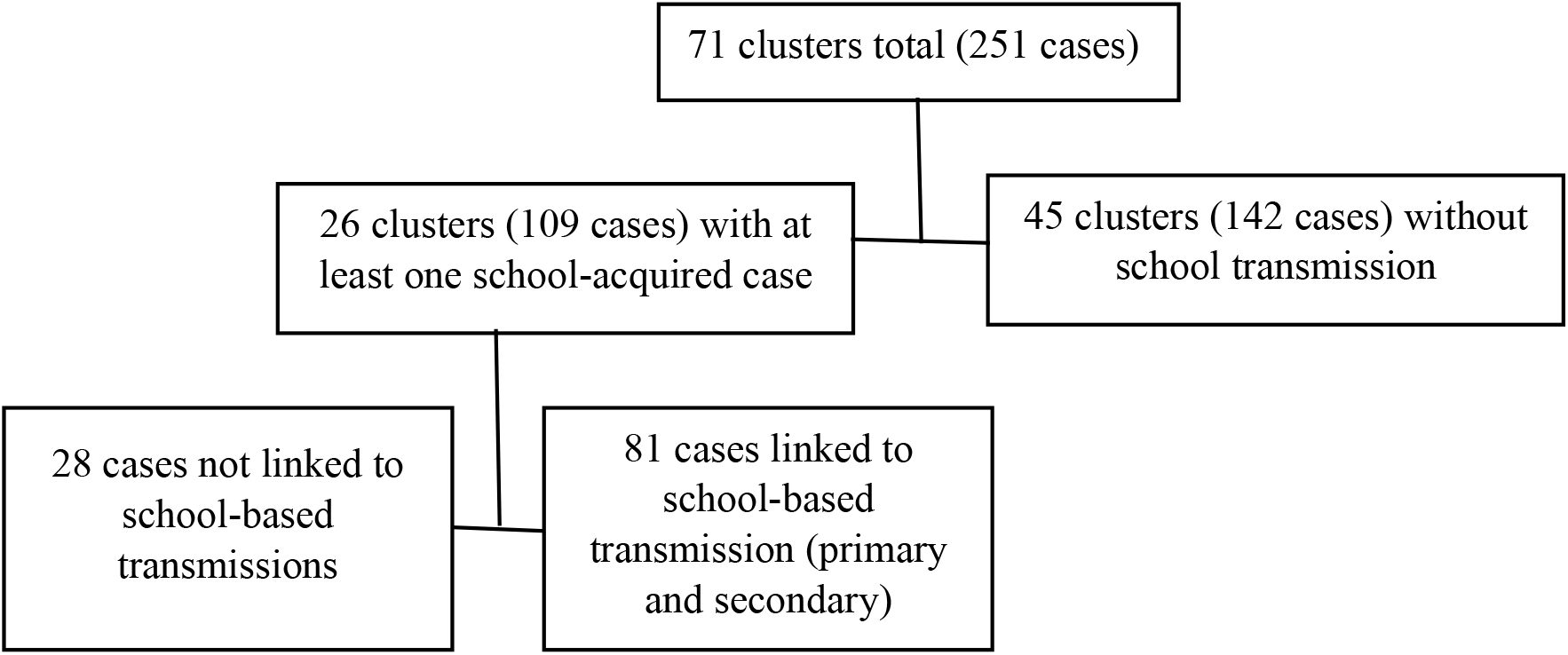
School clusters identification flow diagram.

## Notes

### Competing Interest Statement

The authors have declared no competing interest.

